# A computational analysis on Covid-19 transmission raises imuuno-epidemiology concerns

**DOI:** 10.1101/2020.11.11.20229641

**Authors:** Anthony M. Kyriakopoulos, Shi Zhao

## Abstract

For Severe Acute Respiratory Syndrome Coronavirus-2 (SARS-COV-2) the investigation of the heterogeneity of individual infectiousness becomes important due to the cross reactive immunity of general population. Using a sample of infected population with SARS-COV-2 in close geographical proximity to the initial Severe Advanced Respiratory Syndrome-1 (SARS-1) outbreak, we explored the association between infector’s age and dispersion (or heterogeneity) of individual infectiousness (*k*) in order to investigate the relatedness with the age of an individual’s capability to disperse SARS-COV-2. Interestingly, we find a negative association between *k* and increase of infector’s age. Significantly this becomes more evident for the age group of 20-60 years comparing with the infectors with younger age. This raises important immuno-epidemiology concerns for effectiveness of public health measures to contain the disease.

**One Sentence Summary:** Dispersion of Coronavirus Disease-19 in China differed with age.

## Main Text

The resistance of the “host-donor” defines the efficient transmission of the infectious agents *(1)*. This resistance can be described as the immunity of individuals raised to disable transmission. The previous Severe Acute Respiratory Syndrome-1 (SARS-1) patients, even after 17 years post infection poses a sustained-well-developed, specific T cell memory response against SARS-1 Coronavirus (SARS-1-COV). This specific immunity is intensified according to the increased severity of previous SARS-1 clinical condition *(2)*. These SARS-1 patients whilst all poses a long term cross reactive immunity against SARS-1-COV, also all (to a tested sample of 23/23), poses reacting T cells to the N peptides of SARS-COV-2 *(3)*.Further, the unexposed individuals with no previous history of SARS-1 infection, and negative for Nucleocaspid protein (N) antibodies and neutralizing antibodies of SARS-COV-2, to a 51.35 % (19/37), have also a reactive T cell immunity against SARS-COV-2 N, and non structural proteins (nsp) proteins of SARS-COV-2 (50 % and 50 % respectively) *(3)*. Also a widely distributed T cell cross reactivity (35 % – 60 %) amongst unexposed individuals extends against to the SARS-COV-2 spike (S) protein *(4,5)*.

The specific memory immunity of unexposed individuals is attributed either to immune cross reactions with common flu coronavirus infections *(4,5)* or to the involvement of animal species cross transmitting coronaviruses to human *(3)*. Since there is a 17 year interval from SARS-1 epidemic and the developmental stages of immunity can be severely influenced *(6,7)* by the cross barrier transmission of coronaviruses (including common cold coronaviruses) between animal species and human individuals “Fig. 1”, we have selected specifically to investigate the variation of dispersion parameter *k* with age increase, bearing in mind the concept of higher *k* → less heterogeneity of age group → more difficult to control the epidemic *(8)*. We considered that investigation on this route could identify distinct variation in individual infectiveness with the new virus and thus identify variations on the efficiency of individuals to spread SARS-COV-2 between specific age groups.

**Fig. 1.**
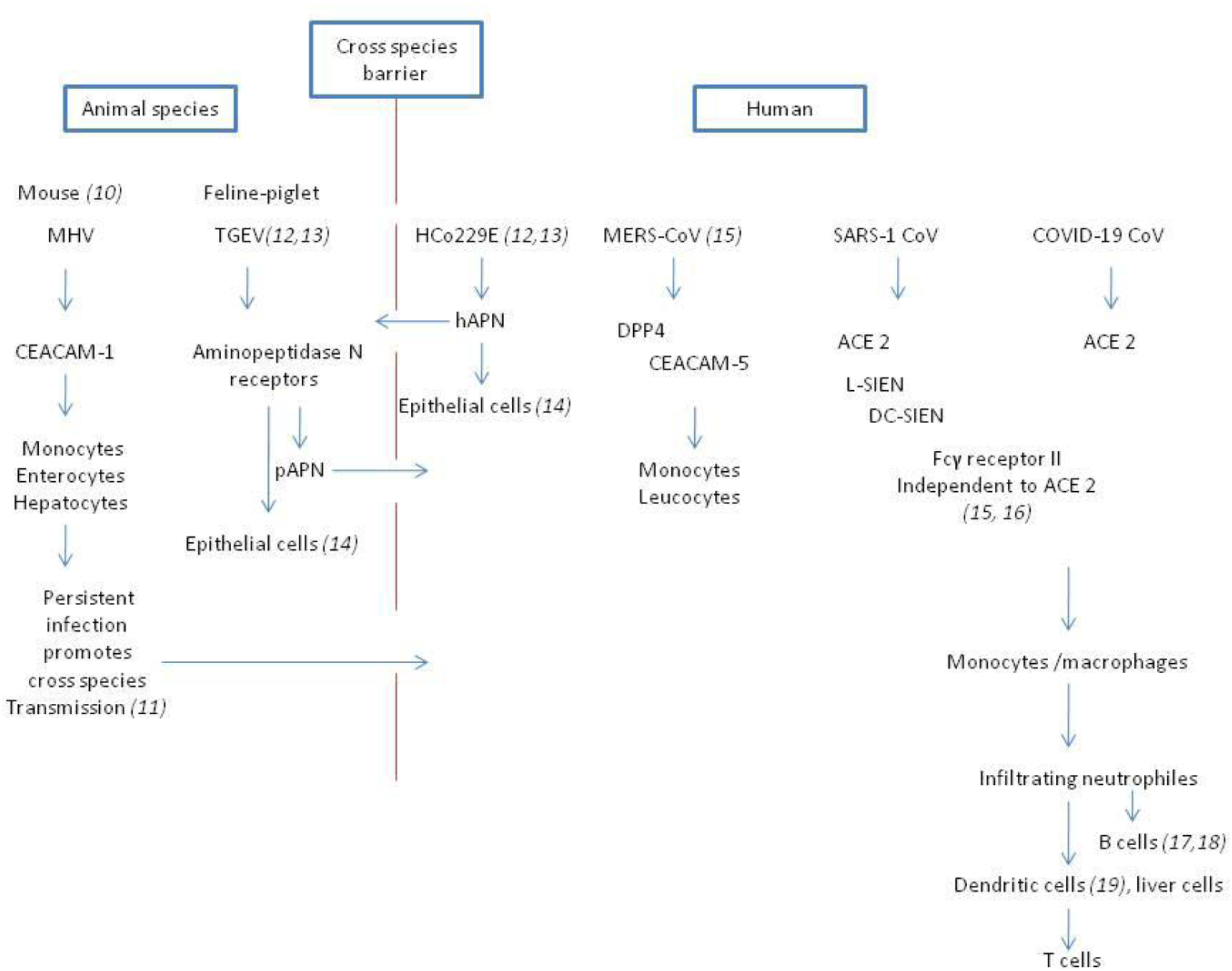
Cross species barrier transmission between human and animals by coronaviruses. The cross species barrier infection is achieved by coronaviruses using suitable receptors that enable this transmission. Molecules like the carcinoembryonic antigen-related adhesion molecules (CEACAM) comprise a family of antigens which are highly preserved between animal and human species and their participation can lead to a wide immune cellular dispersal of coronaviruses throughout the human organism *(9)*.

pAPN: Piglet aminopeptidase N receptor; hAPN: Human aminopeptidase N receptor; L-SIEN: Liver/Lymph node specific intracellular adhesion molecule 3-grabbing integrin; DC-SIEN: Dendritic cell specific intracellular adhesion molecule 3-grabbing non integrin We have therefore employed the metric named dispersion of individual infectiousness, denoted by *k*, which was first proposed for SARS-1 *(20)* since it was used to quantify the role of heterogeneity of individuals in transmitting Coronavirus Disease-19 (Covid-19) as well as the difficulty in controlling the epidemics with Non Pharmaceutical Interventions (NPIs) at population scale *(21,22)*. We performed a statistical calculation of *k*, and explored to define possible relatedness of infector’s age as a determinant of *k*. We estimated the *k* ranges from 0.4 to 1.5 for different age bins, which is in line with previous studies *(8, 21,22)*. We observe an evident downward trend in k as the infector’s age increases, with p-value < 0.001 for linear trends testing using Student’s t test. Moreover, we detect a structural break at age bin 20-40 years, in which k drops 47% with p-value < 0.001 comparing with the infector with younger age “Fig. 2”.

**Fig. 2.**
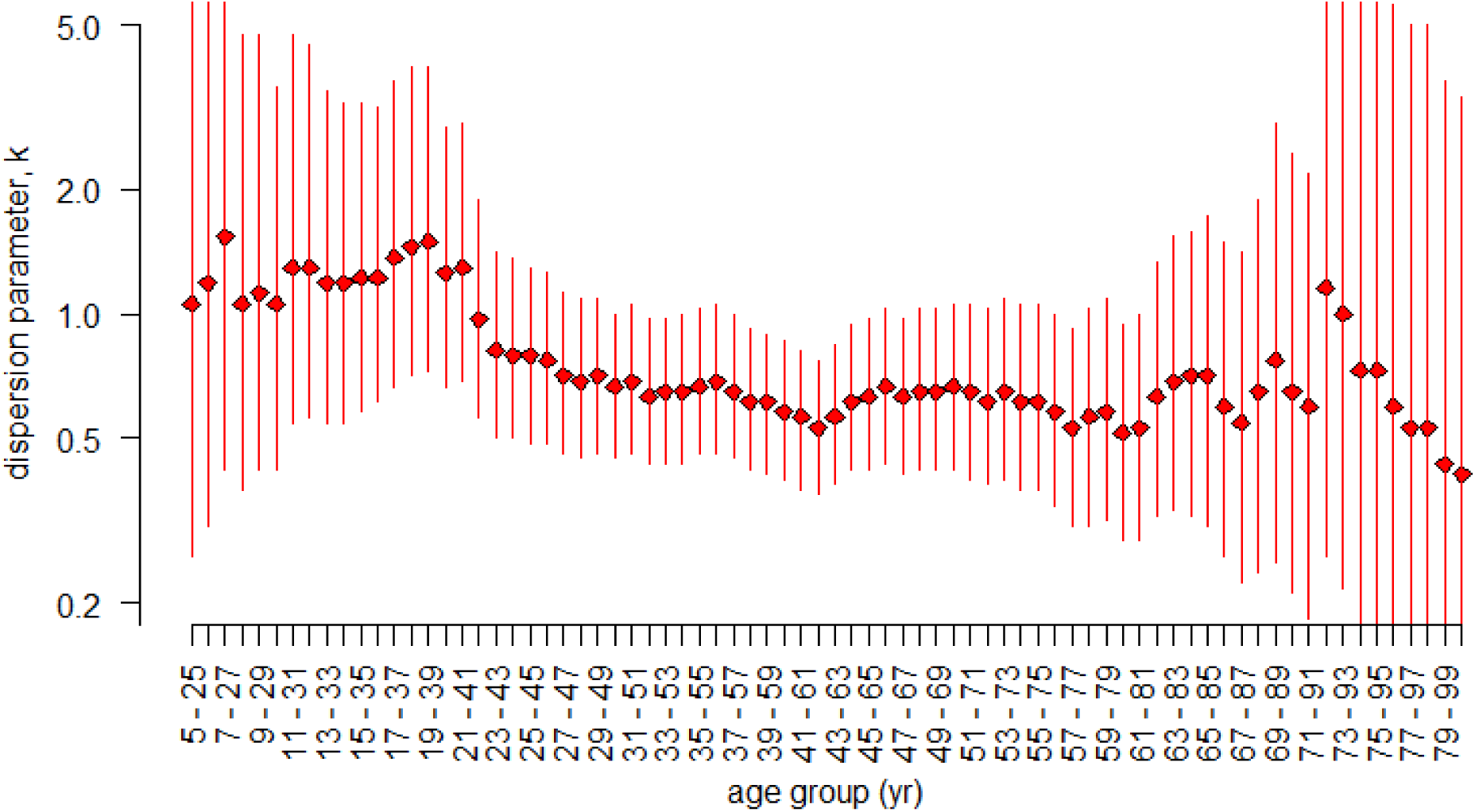
The dispersion parameter (*k*). The estimated dispersion parameter *(k)* in different age sub-groups. The dots are the estimates, and the vertical bars are the 95%CI.

The dataset [see *(23)* for full list of transmission pairs] we used to investigate variation in heterogeity of individual infectiveness with age initially contained 1407 transmission pairs *(22)*, out of which we have identified 807 infectors. Out of these infectors, we selected 777 with age information.

In the investigation of individual infectiveness we kept the reproduction number R constant in order to measure the variation of individual dispersiveness, *k*, with age. The *k* value decreases with age increase, with the important difference being between the two age groups 0-20, and 20-60 years of age (Fig.2). This is important, as their difference lies in the heterogeneity of each sub-population group *(8,20)*. For youth age as *k* is larger, the heterogeneity of subgroup population is smaller and for the older age group as *k*, decreases the heterogeity between individual increases *(20, 22, 24)*. Hypothesizing that R equals to 2, which is a realistic scenario for SARS-COV-2 *(25)*, for four seed cases of youth and old age respectively, the offspring cases will be eight for each age group. This means that almost all youth age infectors will produce two offspring cases, whereas for old age infectors, almost half will not be able to produce any offspring case, one will be able to produce six offspring cases and one two offspring cases “Fig.3”.

**Figure 3.**
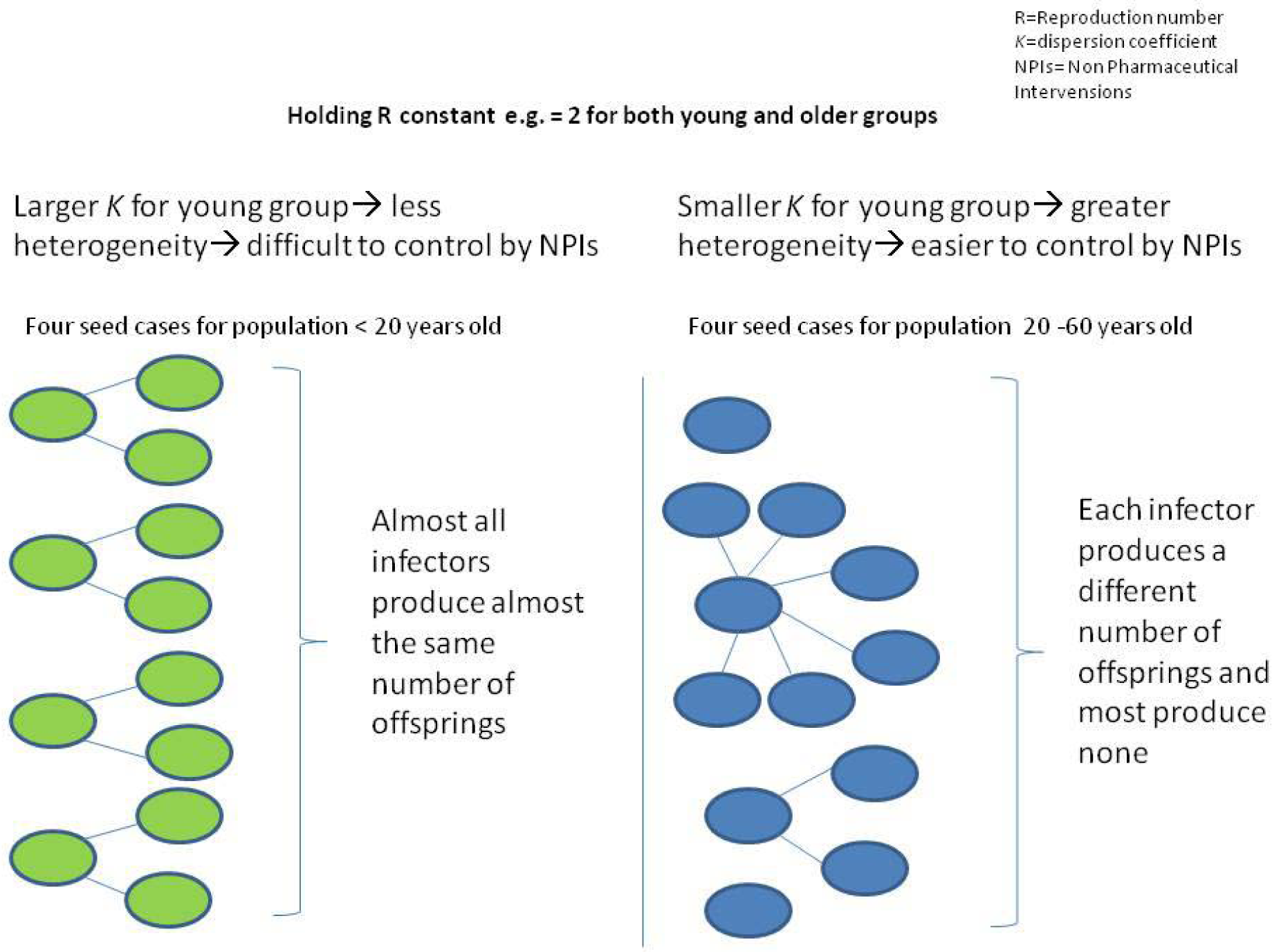
The difference of *k* between age groups concerns the ability of infectors to transmit the disease.

Reflecting the change of individual infectiveness with age and thus their heterogeneity, NPIs, are more applicable to the sub-population older than 20 years of age *(20)*, whereas NPIs are not expected to provide an adequate solution *(26)*, for disease spread containment from younger age infectors. Given the situation that a high proportion of youth age remains asymptomatic but highly infectious *(6, 27)*, this makes the contact tracing effort in this group even more difficult but urgent. Specific screening strategy for the youth population to identify as more possible positivity will make restriction contact measures more efficient as almost all young infected individuals are likely to transmit the disease. The notable differences in the heterogeity of individuals across the age groups of 0-20, 20-60 and over 60 years old may reflect de-similarities between developments of immune surveillance mechanisms due to environmental cross transmission reactions *(10-19)*. These are encountered in previous SARS-1 patients and unexposed individuals *(3-5)*. Due to Covid-19 overspread and consistence, the targeted pharmaceutical interventions may be appropriate to lower seed cases in population groups with small heterogeneity. As our results show, by focusing on youth age population to prevent from spreading SARS-COV-2 to rest of general population, this will help to contain pandemic in a similar way to SARS-1 *(26)*.

## Data Availability

All data is available in the main text or the supplementary materials data file.

## Acknowledgments

We thank Professor J. Kountouras and Professor Markus Nagl for their valuable communication on Covid-19 epidemiology. We thank our families for moral support in conducting this research.

## Funding

This research did not receive any specific grant from funding agencies in the public, commercial, or not-for-profit sectors.

## Author contributions

AK inspired the hypothesis of propagation of Covid-19 through different immune surveillance of population groups and hypothesized for age differences. AK has written the manuscript and SZ conducted the proper mathematical analysis to investigate the AK hypothesis and performed the mathematical part of this manuscript. SZ has written the materials and methods.

## Competing interests

Authors declare no competing interests.

## Data and materials availability

All data is available in the main text or the supplementary materials.

## Supplementary Data for

## Materials

### Data collection and extraction

We used the COVID-19 surveillance data previously published *(23)* and the dataset can be accessed freely via the public respiratory https://github.com/linwangidd/covid19_transmissionPairs_China/blob/master/transmission_pairs_covid_v2.csv.

The dataset contains 1407 transmission pairs that are identified and reconstructed according to the previous studies *(8, 20)*, governmental new release, and official situation reports.

We identified 807 infectors, and who act as source of infection to transmit to the infectee. We extract the information, including age and gender, of each infector as well as the number of offspring infectees generated by each infector. After excluding the infector with missing information on age, we collected 777 infectors for further analysis.

## Methods

### 1. Heterogeneity of individual infectiousness: a statistical modeling perspective

We consider the variation in the individual-level infectiousness as a quantifiable scale that affects the distribution of offspring infectee generated by an infector. Following the previous study *(20)*, we introduce the number of offspring infectee generated by an infector, denoted by *r*, as a random variable from a Gamma distribution, denoted by *h*(), with mean *R* (> 0) and dispersion parameter *k* (> 0). Thus, we have *r* ∼ *h*(*R, k*). Here, *R* is the reproduction number that is defined as the expected (or average) number of secondary cases caused by one typical infected individual.

The dispersion parameter *k* governs the dispersiveness of the Gamma distribution. As demonstrated theoretically in the previous study (*20)*, with *R* fixed, a larger *k* results in a lower effectiveness of non-pharmaceutical interventions in controlling the epidemics, which is also discussed in another study *(8)*.

Poisson process with rate *r*, denoted by *f*(*r*), is adopted to address the stochastic effects in transmission, and to govern the number of infectee caused by each infector, denoted by *Z* (≥ 0). *(29)*. Thus, we have *Z* ∼ *f*(*r*) = *f*(*R, k*). Straightforwardly, *f*(*R, k*) is an Negative Binomial (NB) distribution with mean *R* and variance *R*·(1 + *R*/*k*). By the definition of NB distribution, the probability that one infector generates (*j*−1) offspring infectees, i.e., cluster size of *j* (≥ 1), which is denoted by Pr(*Z* = *j*−1) = *L*_*j*_, is given in Eqn (1).

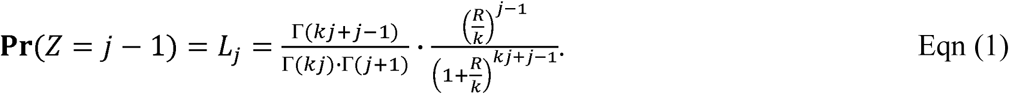

Here, Γ(·) denotes the Gamma function. Specially, the NB distribution *f*(·) reduces to a Geometric distribution when *k* = 1, and it reduces to a Poisson distribution when *k* approaches infinity. Importantly, a smaller value of *k* indicates larger heterogeneity in individual infectiousness.

By fitting distribution *f*(·) to the real-world observations, we may estimate value of dispersion parameter *k*, and explore the determinants of *k*.

### 2. Likelihood inference framework and subgrouping by infector’s age

We consider observed samples of number of offsprings from *N* infectors. We denote the number of infectors who have *j* infectees associated by *n*_*j*_ (≥ 1). Note that all *j* > 1 in our dataset though *j* may be 1 theoretically or observed in other datasets, and thus we adjusted for this truncation in our likelihood framework. Straightforwardly, we have ∑_*j*>1_ *n*_*j*_ = *N*. Then, following the previous studies *(21, 28)*, we constructed the likelihood function, denoted by *L*, as in Eqn (2).

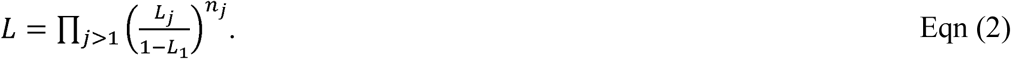

We estimated the dispersion parameter *k* using the maximal likelihood estimation approach. To explore the association between the infector’s age and *k*, we repeated the above fitting and estimation procedure after sub-setting the dataset into subgroups by the infector’s age. We considered 76 age bins, and they include 5-25, 6-26, …, 79-99, and 80+ years. We estimate the value of *k* for each age bin to examine the association between the infector’s age and *k*.

## Supplementary text

### Estimating results

We estimated the *k* ranges from 0.4 to 1.5 for different age bins, which is line with previous studies *(8, 21, 22)*. We observe an evident downward trend in *k* as the infector’s age increases, with *p*-value < 0.001 for linear trends testing using Student’s *t* test. We detect a structural break *(30)* at age bin 20-40 years, in which *k* drops 47% with *p*-value < 0.001 comparing with the infector with younger age seen in “Fig. 2”.

## Notes

### Competing Interest Statement

The authors have declared no competing interest.

### Author Declarations

No special reporting quidelines found according to equator network

